# Rapid Impact Analysis of B 1.1.7 Variant on the Spread of SARS-CoV-2 in North Carolina

**DOI:** 10.1101/2021.02.07.21251291

**Authors:** Michael DeWitt

## Abstract

**Background:** Several cases of the B1.1.7 variant of the SARS-CoV-2 virus were identified in North Carolina first on January 23, 2021 in Mecklenburg County and later in Guilford County on January 28, 2021.^[1,2]^ This variant has been associated with higher levels of transmissibility.^[3–6]^ This study examines the potential impact of increased transmissibility as the B1.1.7 variant becomes more predominant given current vaccine distribution plans and existing non-pharmaceutical interventions (NPIs).

**Method:** We explored the anticipated impact on the effective reproduction number for North Carolina and Guilford County given the date of import of B1.1.7. The approximate growth rate in proportion of B1.1.7 observed in the United Kingdom was fit and used to establish the estimate share of B1.1.7 circulating in North Carolina. Using the nowcasted reproduction numbers, a stochastic discrete compartmental model was fit with the current vaccination rates and B1.1.7 transmissibility to estimate the impact on the effective reproduction number.

**Results:** We found that the effective reproduction number for North Carolina and Guilford County may exceed one, indicating a return to accelerating spread of infection in April as the proportion of B1.1.7 increases. The effective reproduction number will likely decrease into March, then increase as the proportion of B1.1.7 increases in circulation in the population.

**Conclusions:** Existing non-pharmaceutical interventions will need to remain in effect through the spring. Given the current vaccination rate and these interventions, it is likely that there will be an increase in SARS-CoV-2 infections. The impact of the variant will likely be heterogeneous across North Carolina given the reproduction number and volume of susceptible persons in each county at the time of introduction of the variant. Age-based vaccinations will likely reduce the overall impact on hospitalizations. This analysis underlines the need for population level genetic surveillance to confirm the proportion of variants circulating.

## Method

### Data

This analysis considered effective reproduction number data generated for the state of North Carolina using the^[7]^ R package following the methods specified by.^[8]^ Data on S-gene target failure (SGTF) observed in the United Kingdom and made available in the analysis^[5]^ were used in order to estimate proportion of anticipated B1.1.7 variant circulating. Case rates were provided by the North Carolina Department of Health and Human Services and retrieved using the nccovid package.^[9]^

### Statistical analysis

In order to estimate the growth in proportion of B1.1.7, a hierarchical beta regression model was fit to data available from the analysis of Davies^[5]^ representing the proportion of SGTF cases out of cases tested using a specific testing platform within seven NHS England regions. In this model, the outcome variable was the proportion of SGTF given the number of days after introduction. The NHS England region was used as a group level intercept, fit using the RStanArm R package^[10]^. These data were then used to estimate transmissibility multiplier for day t, *ϕ*_*t*_, on the base contact rate given the days after the introduction of the new variant. The transmissibility multiplier represents the weighted average increase in transmission given a fixed contact rate, *β*.

A stochastic discrete compartmental model was then used to simulate the effect of vaccinations and circulation of B1.1.7 on the effective reproduction number. Using case data and including multipliers to account for under-ascertainment of infections, the model included compartments for susceptible, exposed, infected, and removed^1^ persons in the population as shown in the below equations. It is assumed that immunity is permanent for the course of the simulation. The models were fit using the Odin R package^[11]^ and each simulation was run 100 times.

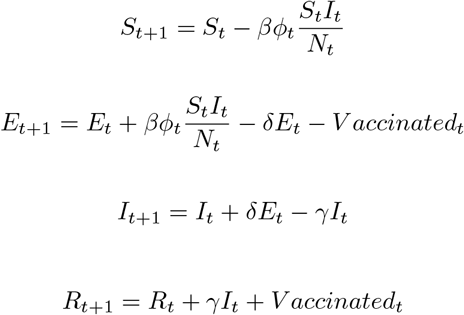

Nine scenarios were evaluated for North Carolina and Guilford County. These scenarios included the 10%, 50%, and 90% quantile estimates for the effective reproduction number on the incubation time adjusted import date of B1.1.7 estimated using the EpiNow2 package.^[7]^ The increase in transmissibility was modeled as 0% reflecting no increase, 50% increase, and 80% increase.^[4,5,14]^ Based on the latest findings from Davies, the increase in transmissibility could be as high as 82% with the 95% credible interval including 106%.^[6]^

The scenarios are shown in Table 1.

**Table 1:** Parameters used in Discrete Stochastic Compartmental Model

| Parameter | Guilford County | North Carolina |
| --- | --- | --- |
| Import Date | 2021-01-28 | 2021-01-23 |
| Reproduction Number | 0.97(0.88-1.03) | 0.96(0.89-1.02) |
| Population | 545,348 | 10,630,691 |
| Susceptible | 454,109 | 8,626,950 |
| Exposed | 1,964 | 31,944 |
| Infected | 3,928 | 63,888 |
| Recovered (Natural Immunity) | 76,490 | 1,669,565 |
| Vaccinated | 8,857 | 238,344 |
| Vaccination Rate (Doses per Day) | 825 | 21,082 |
| Vaccine Efficiency | 95% | 95% |
| Vaccine Uptake | 100% | 100% |
| Variant Transmissibility Increase | 0%, 50%, 80% | 0%, 50%, 80% |

<sup>1</sup>Where “removed” could be through recovery, death, or vaccination.

| Parameter | Guilford County | North Carolina |
| --- | --- | --- |
| Incubation (days) | 6 | 6 |
| Recovery (days) | 10 | 10 |

All analysis was conducted in the R Statistical Computing Environment^[15]^ and is available at https://github.com/conedatascience/sgtf-nc.

## Results

Figure 1 represents the observed proportion of SGTF in the NHS England regions. Additionally, the fitted curve from the beta regression is displayed as a dashed line. This fitted curve was used to estimate the proportion of SGTF in the compartmental models given the introduction date.

**Figure 1:**
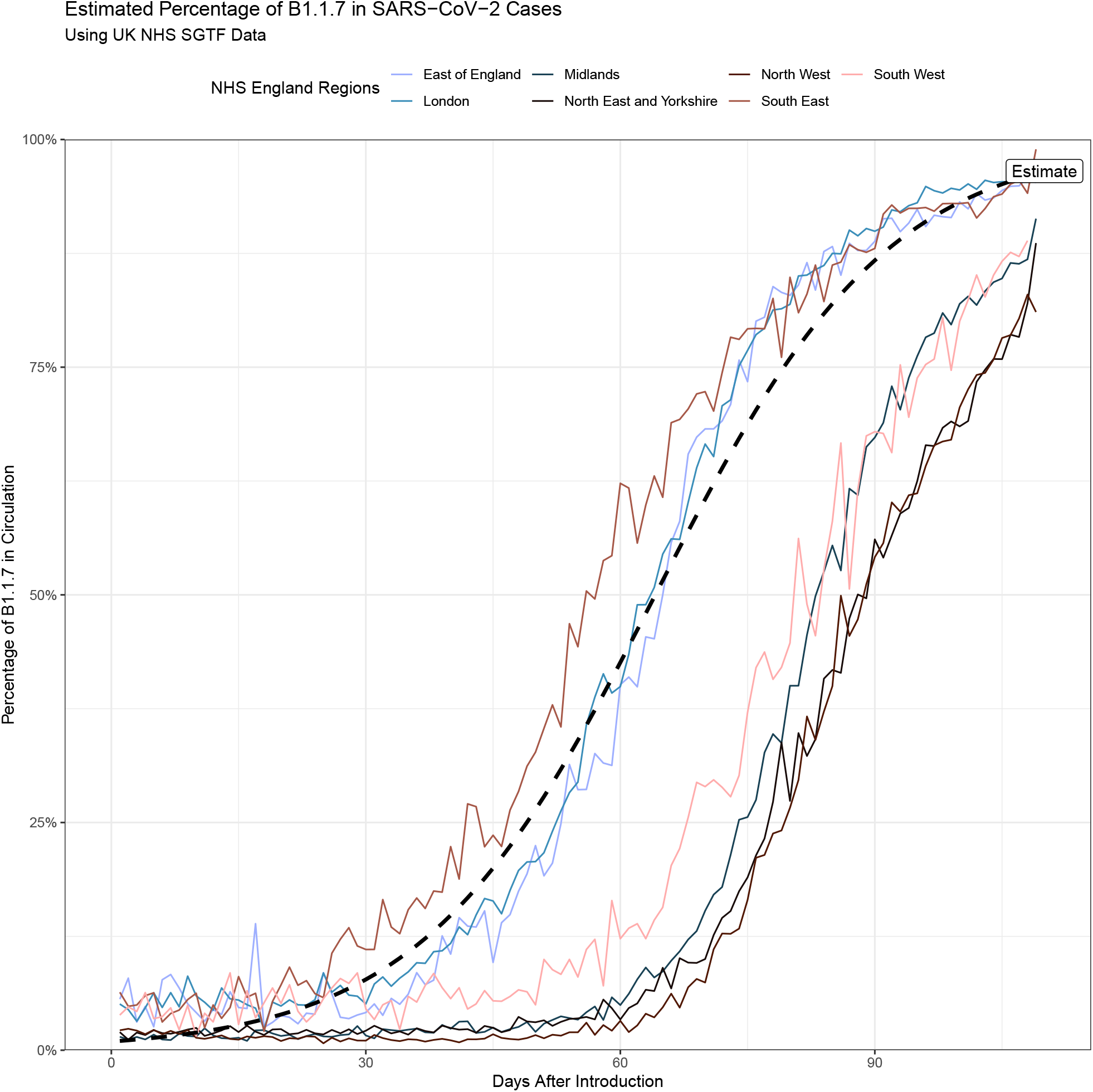
Estimates of SGTF Evolution Used as Proxy for B1.1.7 Share of SARS-CoV-2 Variants in Circulation

Figure 2 shows the evolution in the effective reproduction number in North Carolina and Guilford County.

**Figure 2.**
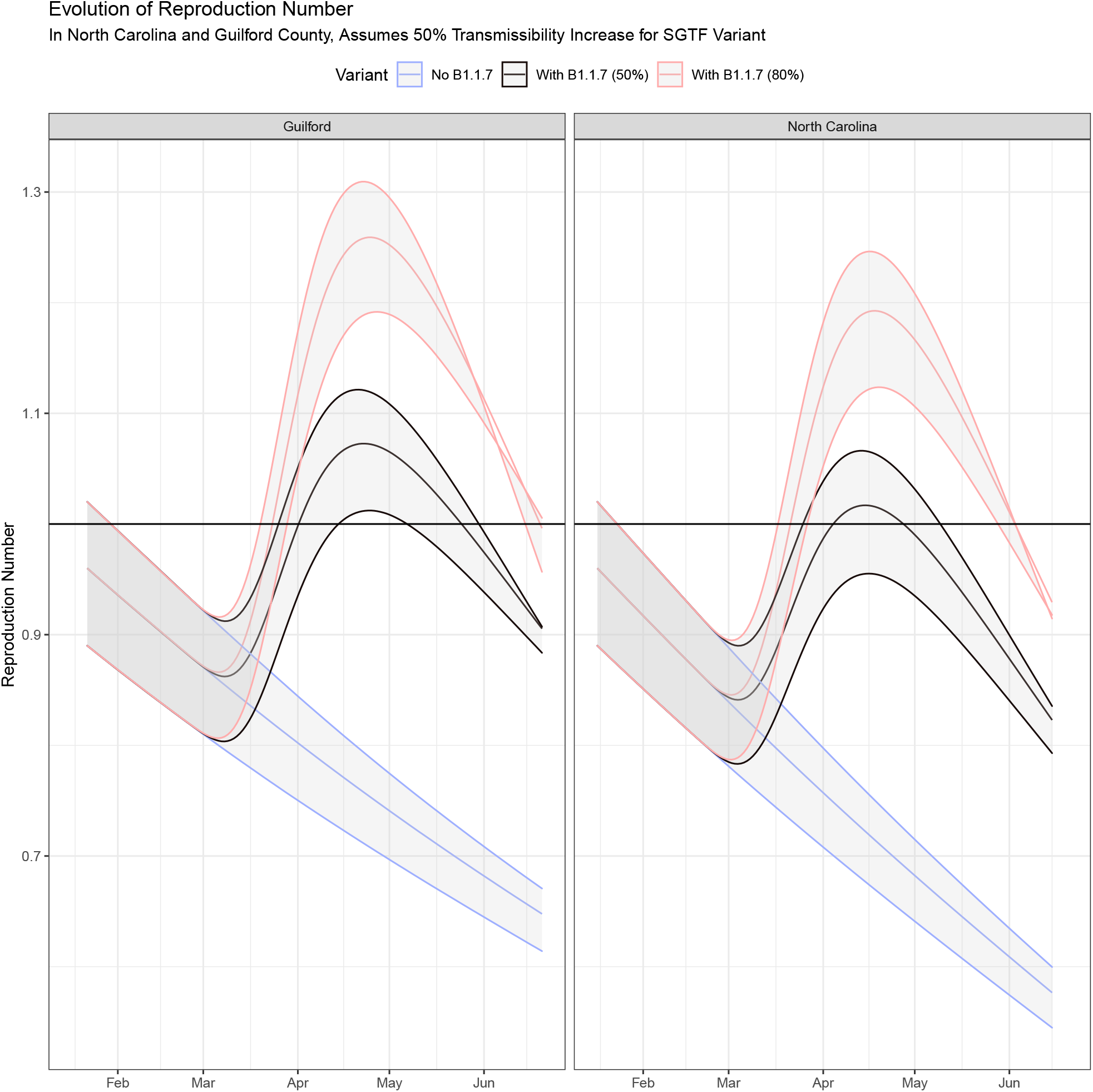
: Estimates of Reproduction Number Evolution in North Carolina and Guilford County Shows Heteogenity

## Discussion

These models demonstrate the potential heterogeneity in impact of the new variant across the state of North Carolina and the importance of the effective reproduction number at the time of introduction of B1.1.7. Both models indicate that if B1.1.7 results in a 50% increase in transmissibility and overtakes existing wild type variants as observed in England, the effective reproduction number may exceed 1, indicating a return to exponential spread in April of 2021. A similar trend would be observed if the increase in transmissibility is as high as 80%. This underlines the importance of accelerating vaccinations across North Carolina. Additionally, this analysis is in the context of existing mobility and occupancy restrictions, indicating that these interventions should remain in effect through the spring.

Importantly, the trajectory for the effective reproduction number will be similar in both regions with and without the variant until March, showing a decrease in the effective reproduction number over the short term. This will be observed in lower incidences of cases. However, as B1.1.7 becomes the dominant variant, the effective reproduction number will likely start to increase. Without rigorous sampling of SARS-CoV-2 infections for genetic sequencing, the proportion of B1.1.7 circulating is unknown. Thus we will not be able to confirm the above analysis until March when the reproduction begins to increase.

This analysis assumes homogeneous mixing in the populations modeled and does not consider differences in contact patterns amongst different age groups. Vaccination uptake is assumed to be 100% over the short term, though as time progresses it may be harder to distribute doses to hard to reach or vaccine hesitant populations. This does not consider the introduction of new vaccine programs, which should increase available supply of vaccine (though at the time of writing only two vaccines have EUA approval from the United States Food and Drug Administration). Similarly, it is estimated the vaccination programs that favor the elderly could have an out-sized impact on the hospitalization rate.^[16,17]^ Additionally, the analysis considers neither the impact of seasonality as this is still an open question, nor the impact of major holiday gatherings like Passover and Easter.^[18]^ It is assumed that existing restrictions on mobility and the requirement for facemasks in public places remains the same.^[19]^ The circulation of other new and emerging variants is not considered.

## Data Availability

All code and data are available at https://github.com/conedatascience/sgtf-nc

## Footnotes

1 Where “removed” could be through recovery, death, or vaccination.

## Notes

### Competing Interest Statement

The authors have declared no competing interest.

## References

1. Department of Health and Human Services, N. C. (2021). NCDHHS: NCDHHS Reports First Identified Case of B.1.1.7 COVID-19 Variant in NC. https://www.ncdhhs.gov/news/press-releases/ncdhhs-reports-first-identified-case-b117-covid-19-variant-nc

2. Guilford County Department of Health. (2021). Guilford County Division of Public Health Confirms First Case of COVID-19 B.1.1.7 Variant in Guilford County Public Health News Guilford County, NC. https://www.guilfordcountync.gov/Home/Components/News/News/2317/1047?backlist=EstimatedPercentageofB1.1.7inSARS-CoV-2Cases

3. Public Health, E. (2021). Investigation of novel SARS-CoV-2 variant - Variant of Concern 202012/01 (Technical Report No. 5; p. 19).

4. Volz, E., Mishra, S., Chand, M., Barrett, JC., Johnson, R., Geidelberg, L., Hinsley, W., Laydon, D., Dabrera, G., O’Toole, A., Amato, R., Manon Ragonnet-Cronin, I., Harrison Jackson, B., Ariani, CV., Boyd, O., Loman, N., McCrone, J., Gonçalves, S., Jorgensen, D., … Ferguson, N. (2020). Transmission of SARS-CoV-2 LineageB.1.1.7 in england: Insights from linkingepidemiological and genetic data. https://www.gov.uk/government/publications/investigation-of-novel-sars-cov-2-variant-variant-of-concern-20201201.

5. Davies, N. G., Barnard, R. C., Jarvis, C. I., Kucharski, A. J., Munday, J., Pearson, C. A. B., Russell, T. W., Tully, D. C., Abbott, S., Gimma, A., Waites, W., Wong, K. L., Zandvoort, K. van, Eggo, R. M., Funk, S., Jit, M., Atkins, K. E., & Edmunds, W. J. (2020). Estimated transmissibility and severity of novel SARS-CoV-2 variant of concern 202012/01 in england. medRxiv. https://doi.org/10.1101/2020.12.24.20248822

6. Davies, N. G., Barnard, R. C., Jarvis, C. I., Kucharski, A. J., Munday, J., Pearson, C. A. B., Russell, T. W., Tully, D. C., Abbott, S., Gimma, A., Waites, W., Wong, K. L., Zandvoort, K. van, CMMID COVID-19 Working Group, Eggo, R. M., Funk, S., Jit, M., Atkins, K. E., & Edmunds, W. J. (2021). Estimated transmissibility and severity of novel SARS-CoV-2 Variant of Concern 202012/01 in England [Preprint]. Epidemiology. https://doi.org/10.1101/2020.12.24.20248822

7. Abbott, S., Hellewell, J., Sherratt, K., Gostic, K., Hickson, J., Badr, H. S., DeWitt, M., Thompson, R., EpiForecasts, & Funk, S. (2020). EpiNow2: Estimate real-time case counts and time-varying epidemiological parameters. https://doi.org/10.5281/zenodo.3957489

8. Abbott, S., Hellewell, J., Thompson, R., Sherratt, K., Gibbs, H., Bosse, N., Munday, J., Meakin, S., Doughty, E., Chun, J., Chan, Y., Finger, F., Campbell, P., Endo, A., Pearson, C., Gimma, A., Russell, T., null, null, Flasche, S., … Funk, S. (2020). Estimating the time-varying reproduction number of SARS-CoV-2 using national and subnational case counts. Wellcome Open Research, 5 (112). https://doi.org/10.12688/wellcomeopenres.16006.2

9. DeWitt, M. (2020). Nccovid: Pull north carolina covid-19 outbreak information parameters. -, -(-), –. https://doi.org/10.5281/zenodo.4157121

10. Goodrich, B., Gabry, J., Ali, I., & Brilleman, S. (2018). Rstanarm: Bayesian applied regression modeling via Stan. http://mc-stan.org/

11. FitzJohn, R. (2019). Odin: ODE generation and integration. https://CRAN.R-project.org/package=odin

12. Cevik, M., Kuppalli, K., Kindrachuk, J., & Peiris, M. (2020). Virology, transmission, and pathogenesis of SARS-CoV-2. BMJ, 371. https://doi.org/10.1136/bmj.m3862

13. Ganyani, T., Kremer, C., Chen, D., Torneri, A., Faes, C., Wallinga, J., & Hens, N. (2020). Estimating the generation interval for coronavirus disease (COVID-19) based on symptom onset data, march 2020. Eurosurveillance, 25 (17). https://doi.org/10.2807/1560-7917.ES.2020.25.17.2000257

14. Galloway, S. E., Paul, P., MacCannell, D. R., Johansson, M. A., Brooks, J. T., MacNeil, A., Slayton, R. B., Tong, S., Silk, B. J., Armstrong, G. L., Biggerstaff, M., & Dugan, V. G. (2021). Emergence of SARS-CoV-2 B.1.1.7 Lineage — United States, December 29, 2020–January 12, 2021. MMWR. Morbidity and Mortality Weekly Report, 70 (3). https://doi.org/10.15585/mmwr.mm7003e2

15. R Core Team. (2019). R: A language and environment for statistical computing. R Foundation for Statistical Computing. https://www.R-project.org/

16. Verity, R., Okell, L. C., Dorigatti, I., Winskill, P., Whittaker, C., Imai, N., Cuomo-Dannenburg, G., Thompson, H., Walker, P. G. T., Fu, H., Dighe, A., Griffin, J. T., Baguelin, M., Bhatia, S., Boonyasiri, A., Cori, A., Cucunubá, Z., FitzJohn, R., Gaythorpe, K., … Ferguson, N. M. (2020). Estimates of the severity of coronavirus disease 2019: A model-based analysis. The Lancet Infectious Diseases, 20 (6), 669–677. https://doi.org/10.1016/S1473-3099(20)30243-7

17. Davies, N. G., Klepac, P., Liu, Y., Prem, K., Jit, M., CMMID COVID-19 working group, & Eggo, R. M. (2020). Age-dependent effects in the transmission and control of COVID-19 epidemics. Nature Medicine, 26 (8), 1205–1211. https://doi.org/10.1038/s41591-020-0962-9

18. Neher, R. A., Dyrdak, R., Druelle, V., Hodcroft, E. B., & Albert, J. (2020). Potential impact of seasonal forcing on a SARS-CoV-2 pandemic. Swiss Medical Weekly, 150 (1112). https://doi.org/10.4414/smw.2020.20224

19. Brauner, J. M., Mindermann, S., Sharma, M., Johnston, D., Salvatier, J., Gavenčiak, T., Stephenson, A. B., Leech, G., Altman, G., Mikulik, V., Norman, A. J., Monrad, J. T., Besiroglu, T., Ge, H., Hartwick, M. A., Teh, Y. W., Chindelevitch, L., Gal, Y., & Kulveit, J. (2020). Inferring the effectiveness of government interventions against COVID-19. Science. https://doi.org/10.1126/science.abd9338

